# Disentangling “Bayesian brain” theories of autism spectrum disorder

**DOI:** 10.1101/2022.02.07.22270242

**Authors:** Maya Schneebeli, Helene Haker, Annia Rüesch, Nicole Zahnd, Stephanie Marino, Gina Paolini, Sandra Iglesias, Frederike H. Petzschner, Klaas Enno Stephan

**Author notes:** **Correspondence to:** Maya Schneebeli, Translational Neuromodeling Unit, Wilfriedstr. 6, 8032 Zürich. joint senior authorship (Frederike H. Petzschner, Klaas Enno Stephan).

## Abstract

**Background:** Bayesian theories of perception have provided a variety of alternative mechanistic explanations for autistic symptoms, including (1) overprecise sensations, (2) imprecise priors, (3) inflexible priors, and (4) altered hierarchical learning. Here, we designed a set of experiments to systematically test predictions from each of the four hypotheses in individuals with autism spectrum disorder (ASD).

**Methods:** Two versions of a two-alternative forced-choice (2AFC) random dot motion task were developed to disentangle the influence of sensory inputs and prior expectations and test implications of the four Bayesian hypotheses. Using a cross-sectional observational study design, behavioural data were obtained from participants with autism spectrum disorder (*N*=47) and a control group (*N*=50). Analyses used mixed effects models and a drift diffusion model.

**Results:** Contrary to the sensory overprecision hypothesis, individuals with ASD did not differ from controls in performance or parameter estimates during the perceptual 2AFC task. When adding cues to the task, individuals with ASD profited less from this prior information than controls, as predicted by the imprecise prior hypothesis. However, individuals with ASD were still able to update their initial priors, contradicting the inflexible prior hypothesis. Finally, in line with the hierarchical learning hypothesis, volatility differentially modulated expectation effects, exerting a significantly smaller effect in the ASD group compared to the control group.

**Conclusion:** Our findings support two mutually compatible alterations of perception in ASD (imprecise priors and, to a lesser degree, altered hierarchical learning). By contrast, they are not compatible with the notions of sensory overprecision and inflexible priors.

## Introduction

Autism has long been described as a condition associated with difficulties in social interactions, repetitive behaviours, and difficulties in dealing with changes.^1,2^ While early theories of autism suggested social deficits as the core of the disorder,^3–5^ subsequent work has proposed that autism may be a primarily perceptual disorder of which social deficits arise as secondary symptoms.^6,7^

Concomitant with this shift, Bayesian models of perception have become an increasingly popular framework for explaining autistic symptoms.^8–11^ The key idea behind these Bayesian accounts is that perception combines current sensory information with *a priori* beliefs (Figure 1A) and represents an act of inference, driven by precision-weighted prediction errors^12^ (Figure 1B). In the context of autism, it was suggested that perception in affected individuals is altered by a shift in precision weights, leading to a domination of sensory inputs relative to prior information and increased weighting of prediction errors, which, in turn, results in overfitting and a lack of generalization. Notably, both increased sensory precision and decreased precision of prior beliefs could lead to this imbalance. However, the exact mechanism that causes this imbalance is being debated.^8,11,13,14^

**Figure 1.**
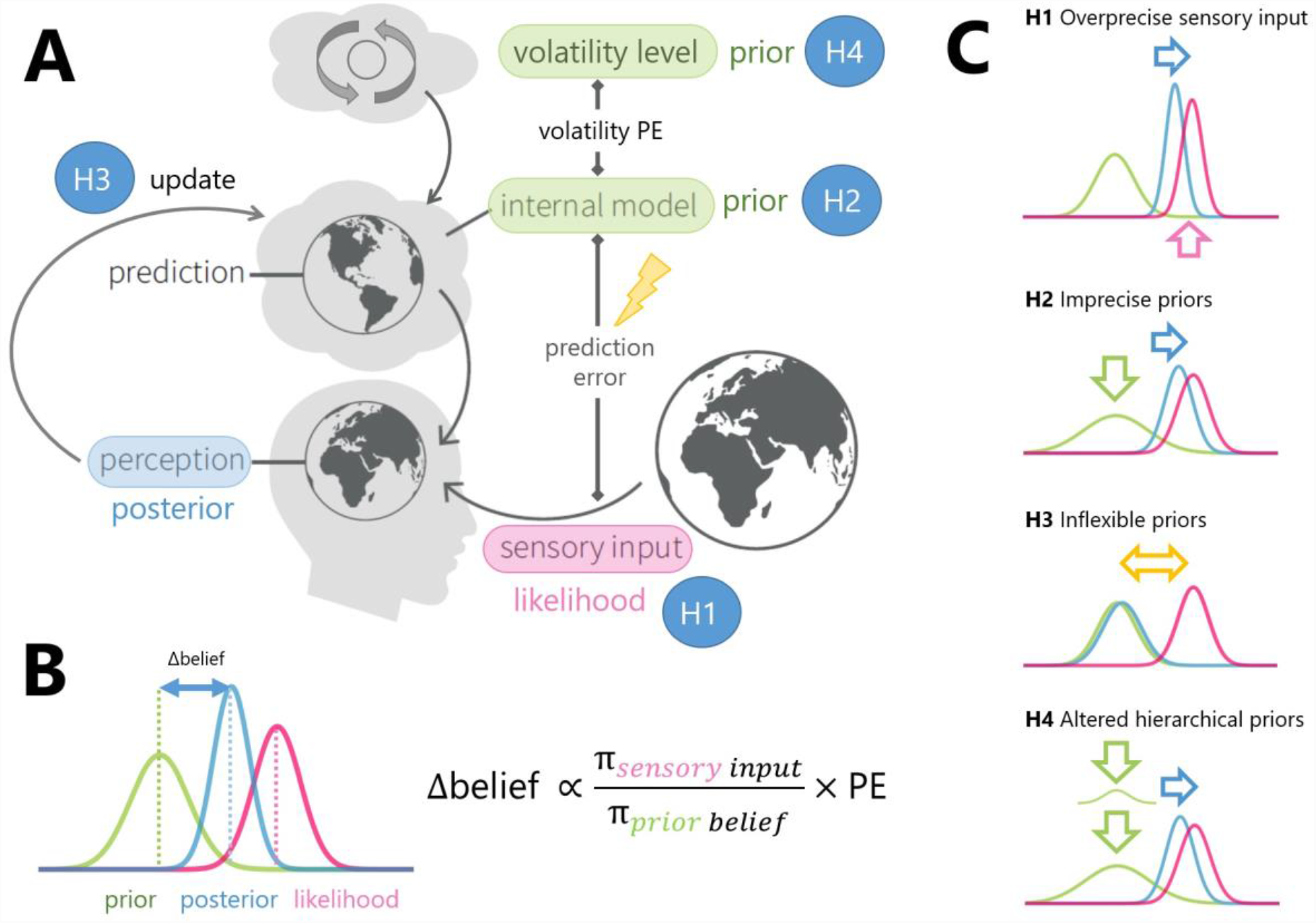
Schematic illustration of a Bayesian model of perception in autism. (**A)** The percept is a combination of the bottom-up sensory information (likelihood) and predictions (prior beliefs) from an internal model of the world. The mismatch between the prediction or prior belief and the sensory input results in a prediction error, which serves to update the belief. (**B)** Left: Illustration of the belief update and percept using Gaussian distributions. Prior and likelihood are combined in a statistical optimal manner (according to Bayes’ rule) to yield a posterior (percept). Right: A generic form of belief updates in Bayesian models: belief updates are proportional to prediction errors, weighted by a ratio of sensory, π_sensory_, and prior precision, π_prior,_. (**C)** Sensory dominance in autistic perception might be caused by different mechanisms: high sensory precision (H1), or low prior precision (H2), high, inflexible prediction errors, indicating a failure of correct prior updating (H3) and deficits in hierarchical learning leading to an overestimation of volatility (H4).

Specifically, there are at least four different mechanisms that could explain an overemphasis of sensory information in a Bayesian framework (Figure 1). As explained in the following, all of these four mechanisms have previously been hypothesized to result in autistic symptoms.

First, individuals with ASD could have more precise sensory channels.^13^ Since the individual weighting of prior expectations in a Bayesian framework depends on the relative uncertainty of the information sources, highly precise sensory information would decrease the effect of prior expectations. This *overprecise sensation hypothesis* (H1) is closely related to the “Enhanced Perceptual Functioning” theory^7^ and motivated by reports that individuals with autism spectrum disorder (ASD) are more sensitive to pitch and loudness in discrimination tasks than healthy controls.^15–17^

Second, sensory processing itself may not be affected in individuals with ASD, but prior information is considered unreliable (i.e. of low precision).^18^ Here, this is referred to as the *imprecise prior hypothesis* (H2).

A third proposal is that individuals with ASD can appropriately acquire prior knowledge but are inflexible in adjusting these beliefs at a later stage. This *inflexible prior hypothesis* (H3) was, for instance, motivated by work showing that individuals with ASD have difficulties updating priors in a multisensory integration task.^19^

Finally, it has been proposed that perceptual deficits in individuals with ASD may arise from abnormalities at higher levels of inference hierarchies. This *hierarchical learning hypothesis* (H4) derives from hierarchical Bayesian models of perception (e.g. predictive coding),^20,21^ where predictions and prediction errors are exchanged between hierarchical layers that represent increasingly abstract aspects of the world. More specifically, this hypothesis suggests that higher processing levels might be more affected than lower ones.^14^ Empirically, this is supported by work showing that in individuals with ASD higher-order beliefs (about volatility) may be overestimated.^22^

While recent studies have begun to test these competing hypotheses empirically,^19,22–35^ the results have been inconsistent and difficult to reconcile. There may be a number of reasons for that: (i) most studies only tested a subsets of the above hypotheses, (ii) some studies did not examine individuals with a diagnosis of ASD, but non-affected individuals with autistic traits ^26,29,31^ and (iii) those that tested diagnosed individuals mostly had relatively small sample sizes, ranging from *N*=14-26.^19,22,24,25,28,30–33,35–37^ Only three studies^23,27,34^ employed larger sample sizes (up to 56) but provided inconsistent results.

Here, we tested predictions of all four hypotheses in a cross-sectional study with 47 participants with ASD (AG: ASD group) and 50 age and gender matched controls (control group; CG). We used two versions of a two-alternative forced choice (2AFC) motion discrimination task in which we probed the integration of sensory processing with prior expectations. The paradigm was specifically designed to disentangle the predictions made by the four hypotheses outlined above and involved several components of learning. Using both model-agnostic analyses and a computational model of the behavioural decision-making, we found evidence supporting the presence of imprecise priors and altered hierarchical learning, but no evidence for the overprecise sensations or inflexible priors hypothesis in ASD.

## Methods

### Study Design

The data presented in this paper are part of a larger study on ASD that was conducted at the Translational Neuromodeling Unit (University of Zurich and ETH Zurich). The study was approved by the cantonal ethics committee (BASEC-Nr: 2016-0838) (see Supplementary Material: Study design).

The analysis plan for this experiment and its revisions are available on an online repository (https://gitlab.ethz.ch/tnu/analysis-plans/schneebelietal_biasd_mdt) (for updates and specifications of the analyses evaluated in the current work, see also Supplementary Material: Analysis plan). A review of the analysis code was conducted by a project member (AR) not involved in the initial data analysis (https://gitlab.ethz.ch/tnu/code/schneebelietal_biasd_mdt). An a priori power analysis (G*Power) indicated an overall sample size of 102 participants (51 per group) to detect a group difference of medium effect size (Cohen’s *d*=0.5, two-sample t-test) with type I error probability of 0.05 and type II error probability of 0.8.

### Participants

To account for possible dropouts, we recruited slightly more participants than the estimated *N*=51 per group. Specifically, 54 participants with a diagnosis of ASD and 57 age-, gender- and handedness-matched controls were included in the study. The diagnosis of ASD was independently verified by a board-certified psychiatrist with specialisation in ASD (HH). After the application of the exclusion criteria for data analysis (see Supplementary Material: Inclusion/Exclusion Criteria), an overall sample of 97 participants remained: 50 controls (CG) (27 male/23 female) and 47 participants with ASD (AG) (17 male/30 female). These sample sizes are slightly smaller than those recommended by the power analysis (i.e., *N*=51 per group). We used the Autism Quotient (AQ) and four neuropsychological tests (see Table 1 and Supplementary Material: Neuropsychological tests) to assess symptom levels and exclude that effects were driven by differences in cognitive and intellectual abilities between the groups.

**Table 1.** Demographics and neuropsychological tests. Table reports mean (standard deviation) or percentage if explicitly mentioned. Significant differences between the control group (CG) and ASD group (AG) were tested for all measures. Levene-Tests were applied to determine equality of variance, determining the test type. As a result, Wilcoxon rank sum tests were applied for AQ, BDT of the HAWIE and age. Sex and handedness were compared using a cross tabulation χ2-test. DSST, d2 and arithmetic scale of the HAWIE was tested with a t-test. Significant differences between groups are indicated by *. Reported effect size (ES) for t-tests is Cohen’s d, for Wilcoxon rank sum test we use effect size r^38^ and for *χ*^*2*^-test we report Cramer’s V. Abbreviations: CG: Control group; AG: autism spectrum disorder group; ES: Effect size; AQ: Autism Quotient; DSST: digital symbol substitution test; BDT: Block design task; d2-R: d2 revised. (See also Supplementary Material: Neuropsychological tests).

| Measure | All (*n*=97) | CG (*n*=50) | AG (*n*=47) |  | *p* | *ES* |
| --- | --- | --- | --- | --- | --- | --- |
| Age (years) | 32.9 (11.7) | 31.5 (10.7) | 34.5 (12.5) | $U=0.91$ | 0.36 | 0.10 |
| Sex (% male) | 0.45 | 0.54 | 0.36 | $\chi^2(1)=3.11$ | 0.08 | 0.18 |
| Handedness (% right) | 0.81 | 0.84 | 0.79 | $\chi^2(1)=0.45$ | 0.50 | 0.07 |
| AQ | 21.5 (12.2) | 13.6 (5.6) | 29.8 (11.7) | $U=6.03$ | <0.001*** | 0.61 |
| DSST | 87.3 (17.6) | 92.5 (14.3) | 81.7 (19.1) | $t_{96}=3.16$ | 0.002** | 0.64 |
| BDT | 51.0 (11.7) | 52.3 (9.8) | 49.6 (13.3) | $U=0.78$ | 0.43 | 0.08 |
| d2-R (F) | 11.8 (17.2) | 12.9 (21.5) | 10.7 (11.1) | $t_{96}=0.63$ | 0.53 | 0.13 |
| Arithmetic scale | 17.1 (3.2) | 17.52 (2.9) | 16.7 (3.4) | $t_{96}=1.24$ | 0.22 | 0.25 |

### Experimental design

The experiment consisted of two consecutive 2AFC tasks: A *non-cued motion discrimination task (NCMDT*; Figure 2A) and a *cued motion discrimination task* (*CMDT*; Figure 2C). In the NCMDT, participants were asked to make decisions about the motion direction of a moving dot stimulus (motion coherence levels in percent: [-51.2, -12.6, -3.2, ±0, +3.2, +12.6, +51.2]%, uniform distribution) (Figure 2B). Responses were made by a saccade towards a Pac-Man shaped target on the left or on the right side and recorded with an infrared eye tracker^39^ (see Supplementary Material: Apparatus). Feedback was given for correct and incorrect responses and a progress bar indicate the accumulated points, which would result in an extra monetary compensation. The CMDT, was identical to the NCMDT except that an arrow cue pointing towards the left or the right side appeared prior to the motion stimulus, which would provide additional prior information about the upcoming motion direction (Figure 2C). Critically, the validity of the cue followed a probabilistic pattern with changing volatility (Figure 2D&E). Additionally, to assess beliefs about the cue in the absence of motion, on every eleventh trial (24 trials) instead of the motion stimulus a question appeared immediately after the cue asking about the expected motion direction and their confidence thereof (Figure 2F). This response was given by pressing the arrows on the keyboard and moving a dot along a line. Overall, participants played 280 trials of the NCMDT and 266 trials of the CMDT in a consecutive order. For detailed descriptions of the tasks see Supplementary Material: Task design.

**Figure 2.**
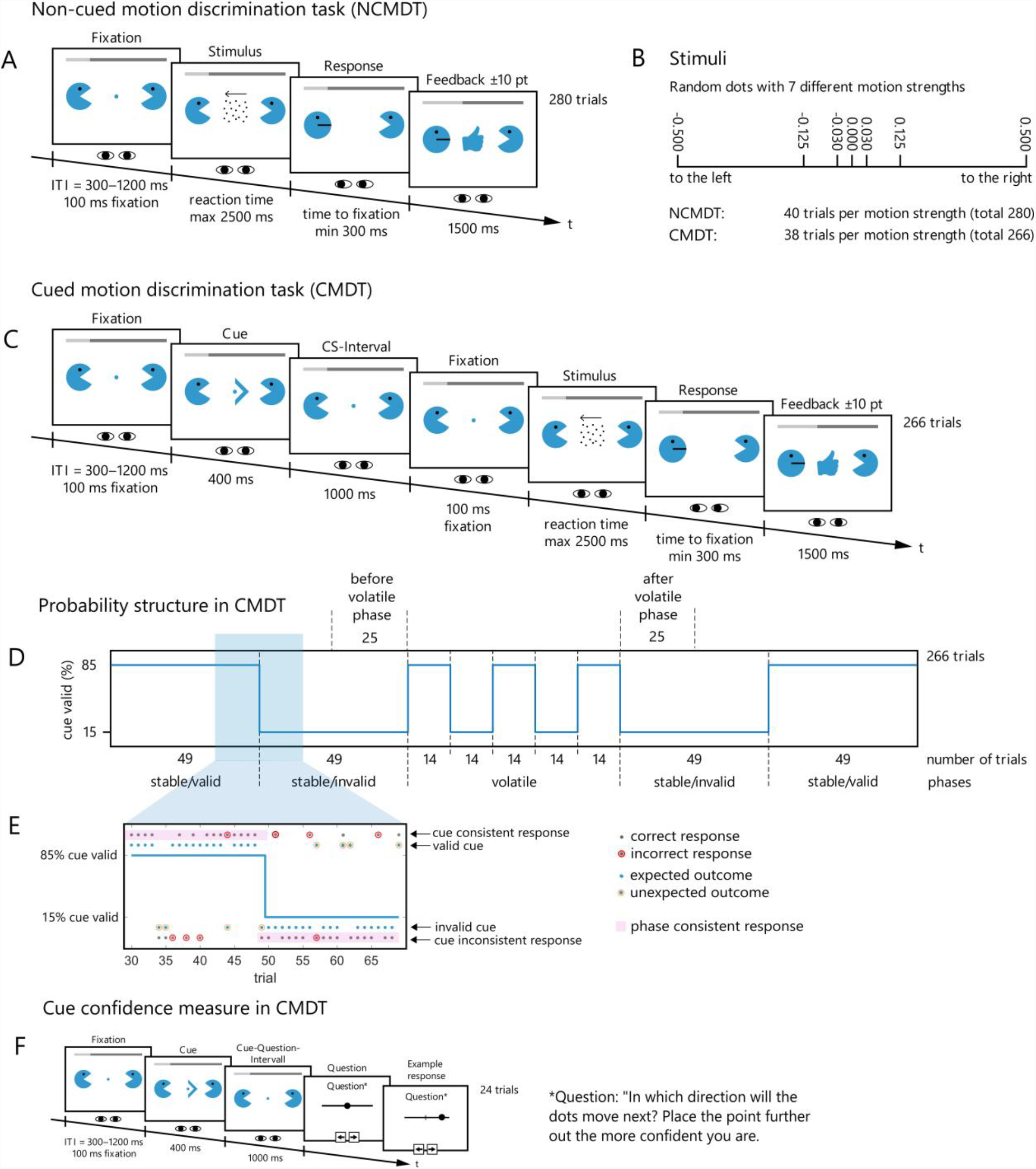
Schematic depiction of the non-cued motion discrimination task (NCMDT) and the cued motion discrimination task (CMDT). (**A)** During the NCMDT, participants were first asked to fixate on the centre of the screen (fixation period). After the inter-trial interval and a successful fixation period of 100ms was registered (eye tracking), the random dot motion stimulus would appear at the centre of the screen until participants indicated their estimated motion with a saccade to the left or right target location (Pac-Man symbol). If no response was given within 2500ms, the trial was registered as a missed trial and feedback was given accordingly. Following a successfully registered saccade, the Pac-Man symbol in the target location would close its mouth. Feedback was shown once participants returned to fixating on the centre again, before the next trial would be initiated. Participants played through 280 trials of the NCMDT. **(B)** One of seven motion strengths was selected for the stimulus in a certain trial in both tasks. The absolute values of the motion strength indicate the fraction of the dots moving accordingly in one direction (negative numbers=leftward motion, positive numbers=rightward motion). **(C)** The trial in CMDT is identical to the NCMDT except that an arrow cue was presented before each stimulus providing additional information about the upcoming motion direction, followed by a cue-stimulus interval (CS-Interval). **(D)** The probability of the validity of the cue varied across the CMDT (valid and invalid phases: 85% and 15% correct indication of the upcoming motion direction, respectively). Moreover, we distinguish between rapid alternations between the two phases (volatile) or longer periods of one validity phase (stable). **(E)** Zoom into a partial sequence of the trace with example responses from a participant. Trials can be separated into different categories (grey dots: response, blue dots: cue). 1. Valid or invalid cue: A valid cue in a trial correctly indicates the upcoming motion direction independently of its phase (upper blue dots). 2. Expected and unexpected outcomes: If a valid cue appears during an invalid phase or an invalid cue appears during a valid phase, the outcome is labelled unexpected (yellow circle around blue dot). 3. Correct or incorrect response: A participant’s response is correct if it indicates the correct motion direction, independently of the cue or phase (red circle around incorrect responses). 4. Cue consistent or inconsistent: If a participant’s response follows the cue independently of its validity, it is labelled as “cue consistent” (upper dots). 5. Phase consistent: If a participant’s response follows the overall phase of the cue (following the cue during valid phases, not following the cue during invalid phases) it is labelled “phase consistent” (pink shaded responses). **(F)** In addition, in 15 trials instead of the stimulus presentation, a question was presented after the cue. Participants were asked to indicate the expected upcoming stimulus direction and their confidence in this prediction.

### Drift-diffusion Model

A drift-diffusion model (DDM) was fit to the choice behaviour and reaction time (RT)^40,41^ (see Supplementary Material: Drift-Diffusion Model). The model used here has three free parameters: a non-decision time *T*_*er*_, a drift *v*, and a boundary *b*. To assess the NCMDT, the starting point *z*_0_ was fixed to 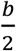, the center between the two boundaries thus indicating no bias towards either choice (left or right) (H1). For the CMDT, the starting point was a free variable which indicated a bias towards the current (*z*_0_ in valid phases and −*z*_0_ in invalid phases). The non-decision time reflects additional sensory and motor delays outside the evidence accumulation process. The drift and the boundary describe the evidence accumulation process. The faster evidence is accumulated, the higher the drift. The boundary describes the threshold, at which point the accumulation process stops and a decision is made. Before fitting the model to our data, we first assured in simulations that all model parameters could be successfully recovered (see Supplementary Material: Parameter recovery analysis).

### Behavioural readouts

For the model-based (DDM) analysis in both the CMDT and NCMDT, we used the percent correct responses (PC) and the reaction time (RT).

In addition, for the model-agnostic analyses, we labelled trials and responses based on the information provided by the cue and the individual response to the cue in the CMDT (Figure 2F). Specifically, we defined the following behavioural readouts:

- Cue consistency: *Cue consistent* trials are trials on which the individual response follows the direction indicated by the current cue. Specifically, during invalid phases, this is a metric which indicates to what extent individuals follow a-priori information provided by the cue itself over the accurately learned phase about the validity of the cue.
- Phase consistency: *Phase consistent* trials are trials on which the judgement of motion direction mirrors the cue during valid phases and opposes to the cue direction during invalid phases. This information is particularly meaningful for zero coherence trials where behaviour is predominantly guided by the cue.
- Outcome expectancy: If a valid cue appears during an invalid phase or an invalid cue appears during a valid phase, the trial outcome is labelled *unexpected*. For instance, during a valid phase of the cue (85% correct predictions of the motion direction), a trial outcome would be considered unexpected if the cue points in the opposite direction of the following motion direction (15% of trials) and conversely for the other (Figure 2E). Note that expectancy here refers to the true probability structure, not to an individual’s trial-wise level of surprise.
- Stability: *Stable* trials include all trials during a stable phase, independently of its validity (first and last 98 trials; Figure 2D). *Volatile* trials correspond to all trials during the volatile phase where the cue validity changes every 14 trials.

In the intermittent question trials (see Figure 2F), a cue was shown without a subsequent stimulus, followed by an explicit question about cue confidence. That is, participants were asked to indicate which motion direction they expected given the cue they just saw. This response was given on a continuous scale which allowed for assessing the confidence in this expectation (compare Figure 2F). The following three definitions are related to these question trials only:

- Reported cue consistency: If participants choose the same direction as the cue, their explicitly reported cue consistency is 1, otherwise it is 0.
- Reported phase consistency: If participants’ explicit prediction was *phase consistent* (see above), the reported phase consistency is 1. Note that this judgment is made in the absence of any stimulus and can thus only rely on the information associated with the cue.
- Reported cue confidence: Absolute distance from 0.5 (centre of the bar). Any value between 0 and 1 was possible (0.01 step size).

### Hypotheses Testing

As described in the Introduction, there are multiple competing Bayesian hypotheses explaining symptoms in ASD: the *overprecise sensation hypothesis* (H1), the *imprecise prior hypothesis* (H2), the *inflexible prior hypothesis* (H3) and the *hierarchical learning hypothesis* (H4). We aimed to disentangle these different hypotheses using a new experimental design and a predefined analysis (the analysis plan incl. its revisions can be found at https://gitlab.ethz.ch/tnu/analysis-plans/biasd_sensory_uncertainty_task). A detailed description of the hypotheses and how we tested them is given in the Supplementary Material: Hypotheses testing. An overview of the tested and confirmed hypotheses is given in Table 3.

For lack of space, the preprocessing procedures used in this study are described in the Supplementary Material: Outlier removal.

### Statistical analyses

In general, we adopted a significance level of *p* ≤ 0.05. For tests between two samples we used the MATLAB (MathWorks, version 2016b) functions *ttest2* for parametric testing of homoscedastic data (tested by Levene’s test), or *ranksum* (Wilcoxon rank sum test) for non-parametric testing and *crosstab* (χ^2^-test) for testing proportions, respectively. Generalized linear mixed effects models (GLMMs) and repeated measures ANOVAs were calculated in R (version 3.5.2) using the package *afex*. Greenhouse-Geissner (GG) correction for the analyses of variance was applied if variances were not equal (tested by Mauchly’s test of sphericity) to reduce type-I-errors. For binary data in mixed effect models, i.e. for the analysis of interaction effects on correct and incorrect trials, we employed a GLMM, therefore GG correction was not applied.

We corrected post-hoc tests within an ANOVA using the family-wise error correction Tukey’s HSD. For stimuli different from zero coherence, we used accuracy and RT as dependent variables. For stimuli with zero coherence, there is no true correct response; therefore, we used phase consistency and cue consistency as dependent variables.

### Data and Code availability

The analysis code can be accessed on the GitLab repository of ETH Zurich (https://gitlab.ethz.ch/tnu/code/schneebelietal_biasd_mdt). Upon acceptance in a peer-reviewed journal, the anonymized data will be made available in the ETH Research Collection (https://www.research-collection.ethz.ch/), a freely accessible scientific repository that complies with the FAIR principles.^42^

## Results

### Demographics and neuropsychology

Between the two groups there were no significant differences in age (Wilcoxon rank-sum test, *U*=0.91, *p*=0.36), handedness (*χ*^*2*^*(1)*=0.45, *p*=0.50), or gender (*χ*^*2*^*(1)*=3.11, *p*=0.08), but a significant difference in the autism quotient (*U*=6.03, *p*<0.001) (Table 1). Furthermore, there were no significant between-group differences in general neuropsychological measures like perceptual logical thinking (BDT: *U*=0.78, *p*=0.43), working memory (arithmetic: t_96_=1.29, *p*=0.22) and attention (d2-R: *t*_*96*_=0.63, *p*=0.53), except for the measure for processing speed (DSST, *t*_*96*_=3.16, *p*=0.002) (Table 1).

### Testing alternatives hypotheses of perceptual alterations in ASD

In the following, we present the results for the hypotheses tested within this study. A detailed description of the hypotheses and the methods used to test them is given in the Supplementary Material: Hypotheses testing.

### Lack of evidence for overprecise sensory information processing in individuals with ASD (H1)

We used a classical 2-AFC motion discrimination task without cue (NCMDT) to capture differences in sensory evidence integration and decision-making during motion discrimination between the AG and CG. We found no significant group differences in average performance and RT (H1a) (Table 2, Figure 3A-C, see also Supplementary Material: Hypotheses testing). Furthermore, the DDM analysis did not reveal any group differences in the boundary and non-decision time parameter estimates. Although there was a marginal difference in the drift parameter (*t*_*95*_=2.04, *p*=0.04; not surviving multiple comparison correction), this effect was in the opposite direction to what would be expected if information processing in the AG were characterised by increased sensory precision: that is, the AG showed a smaller drift compared to the control group. (H1b) (Table 2). This suggests that the AG integrated sensory evidence similarly or even slightly slower than the CG in the NCMDT, and that both groups employed similar decision thresholds.

**Table 2.** Behavioural and model-based summary statistics of the non-cued motion discrimination task. Abbreviations: AG: autism spectrum disorder group, CG: control group, RT: Reaction time (in seconds), PC: Percent Correct responses, b: boundary, v: drift, T_er_: non-decision time in milliseconds. We used t-tests for all comparisons with *df*=96. The last column shows the effect sizes Cohen’s d.

| | | All ($n=97$) | CG ($n=50$) | AG ($n=47$) | $t_{96}$ | $p$ | $d$ |
| --- | --- | --- | --- | --- | --- | --- | --- |
| Behavioural readouts | RT [ms] | 1040 (192) | 1039 (195) | 1041 (191) | 0.07 | 0.94 | 0.01 |
|  | PC [%] | 76.7 (6.48) | 77.2 (6.02) | 76.1 (6.95) | 0.79 | 0.42 | 0.16 |
| Model parameter estimates | b | 1.88 (0.32) | 1.88 (0.34) | 1.89 (0.30) | 0.17 | 0.87 | 0.03 |
|  | v | 7.29 (2.50) | 7.79 (2.22) | 6.68 (2.69) | 2.04 | 0.04 | 0.41 |
| | $T_{er}$ [ms] | 393 (87.4) | 409 (63.6) | 376 (105) | 1.89 | 0.06 | 0.38 |

**Figure 3.**
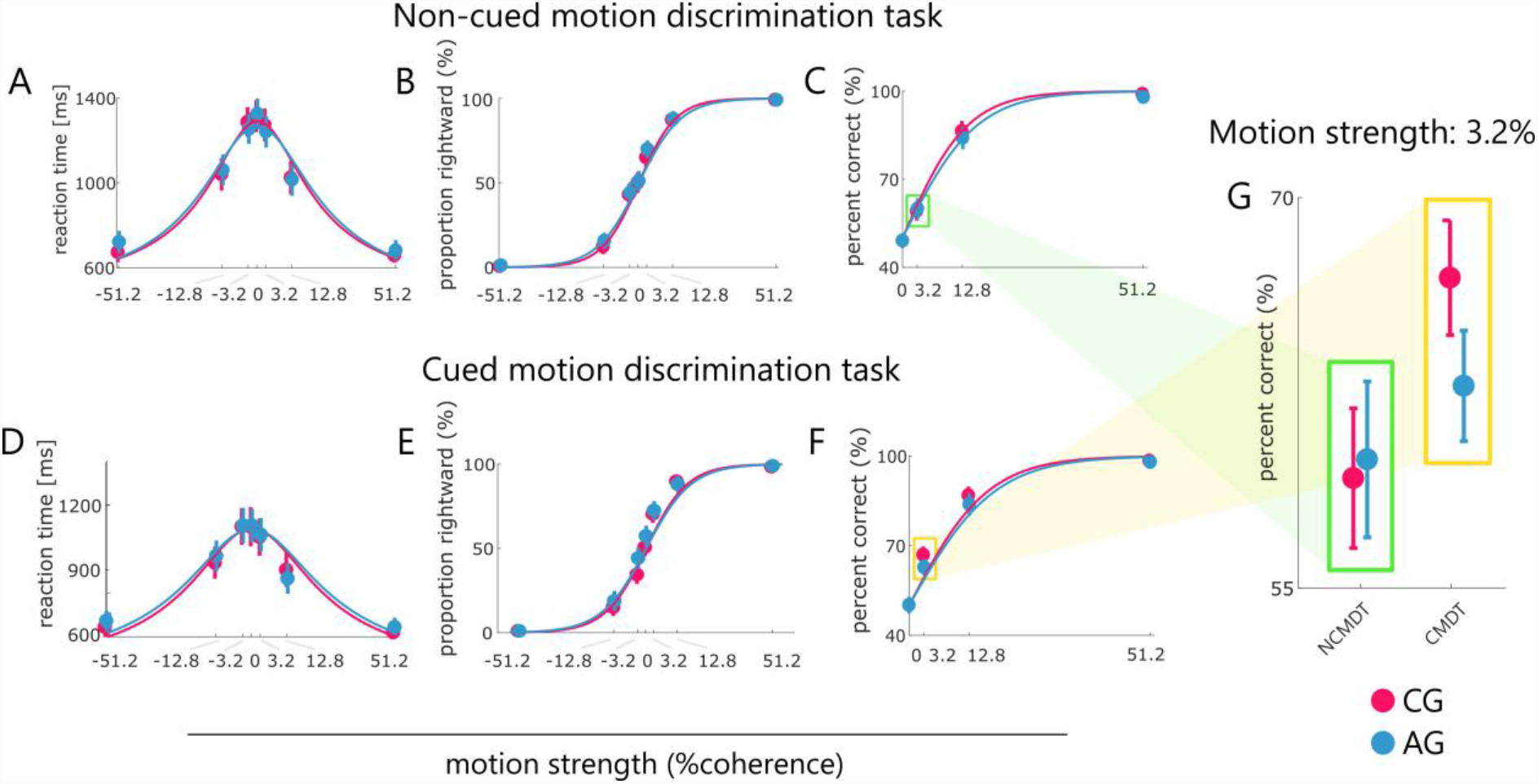
Group differences between tasks (CMDT and NCMDT). Behavioural data (dots) and model predictions (solid line) for the non-cued (upper) and cued (lower) version of the motion discrimination task for both groups (AG in blue, CG in magenta). Model predictions (solid lines) were generated by the DDM, using the empirically obtained parameter estimates and simulating responses across different coherence levels. From left to right: **(A)** and **(D)** Reaction time, **(B)** and **(E)** percent rightward responses and **(C)** and **(F)** proportion correct responses plotted across motion strength levels. In panels (C) and (F), motion strength for left and right was combined and, since there is no correct response for zero coherence stimuli, we assigned a random response (i.e. 50% chance of responding correctly/incorrectly) to these stimuli. Panel **(G)** shows a zoom-in of the highlighted section in (C) and (F) for the 3.2% coherence level across both tasks. Error bars reflect 95% confidence interval. CMDT: cued motion discrimination task; NCMDT: non-cued motion discrimination task; AG: autism spectrum disorder group, CG: control group.

### Differences in the use of prior information (H2)

To assess differences in the use of prior information we used a second paradigm where an initial probabilistic cue indicated the potential motion direction on each trial (CMDT) (Figure 2C). To further assess the effect of learning and hierarchical modulation of prior information the validity of this cue was changed over the course of trials (valid: 85% correct; invalid: 15% correct) and the frequency of these changes was altered (stable vs. volatile phases) (Figure 2E).

### Individuals with autism spectrum disorder use prior information less to make difficult perceptual decisions (H2a)

According to Hypothesis 2, individuals with ASD treat prior information as unreliable (i.e. of low precision). They should thus benefit less from information provided by the cue, and this should become particularly apparent when sensory uncertainty is high.

To test this, we used a mixed effects model for accuracy and RT data, with the fixed factors task (NCMDT vs CMDT) and group (CG vs. AG) and a subject-specific random factor. We found that the task had a main effect on performance (*χ*^*2*^*(1)*=6.9, *p*=0.009). This effect was driven by the CG only (task × group interaction: *χ*^*2*^*(1)*=8.3, *p*=0.004). In other words, while the CG showed a significant increase in performance on the CMDT task, compared to NCMDT (*b*=-0.152 (95%CI [-0.228, -0.076]), *z*=-3.97, *p*=8.6e-5), the AG did not show any benefit of cue (*b*=0.007 (95%CI [-0.070, 0.084]), *z*=0.18, *p*=0.860). We also found a main effect of task on RTs (*F*_*1,95*_=47.5, 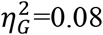, *p*=6.1e-19), but no group × task interaction (*F*_*1,95*_=0.04, 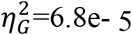, *p*=0.845) (Figure 3A&D).

Next, we tested if the beneficial effects of prior information in the CG (but not AG) was mainly driven by the fact that controls benefit more from the cue when stimuli were difficult and sensory uncertainty was high (i.e. low-coherence trials). This analysis used a GLMM with task, group and difficulty level (coherence) as fixed factors. As expected, there was a highly significant main effect of coherence on accuracy *(χ*^*2*^*(1)*=6493, *p*<2.2e-16) and a task × coherence interaction (*χ*^*2*^*(2)*=18.65, *p*=8.9e-5), indicating that performance increase across tasks differed across coherence levels. Additionally, there was also a marginally significant 3-way interaction effect of task × group × coherence (*χ*^*2*^*(2)*=6.11, *p*=0.047) on accuracy. Tukey-corrected post-hoc analysis revealed that the 3-way interaction effect was driven by the lowest (non-random) motion strength in the cued task between groups. In other words, as predicted by H2, the CG benefited from using the cue on the most difficult (3.2% coherence) trials (*b*=-0.310, 95%CI=[-0.413,-0.207], *z*=-5.88, *p*=2.5e-8) whereas the AG did not (b=-0.050, 95%CI=[-0.156,0.057], *z*=-0.908, *p*=0.800); see Figure 3G. This difference was not found for easier trials where sensory information alone might have been sufficient to provide a correct response (Figure 3F).

### Individuals with ASD are less sensitive to expectations mediated by the cue (H2b)

Another way to test hypothesis 2 is to assess how much the behaviour of both groups is modulated by cue validity. Given the informative nature of the probabilistic cues (85% and 15% cue validity, respectively), task performance and RT can be improved by exploiting the information of the cue about the most probable motion direction. However, due to the probabilistic nature of the cue, this use of prior information makes errors more likely when cue validity is not as expected during the current phase of the task (i.e., a valid cue during an invalid phase and an invalid cue during a valid phase). Indeed overall performance was higher and RT were lower for expected compared to unexpected trial outcomes (GLMM with fixed effects: group and expectancy, random effects: subject; main effect of expectancy on RT: *F*_*1,95*_=61.91, 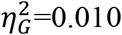, *p*=5.7e-12 and GLMM: main effect of expectancy on accuracy: *χ*^*2*^*(1)*=411.81, *p*=1.5e-91). Across groups, expectancy had a differential influence on accuracy (expectancy × group interaction: *χ*^*2*^*(1)*=9.08, *p*=0.003) but not on RT (expectancy × group interaction: *F*_*1,95*_=0.51, 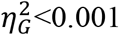, *p*=0.475). This effect was driven by a selective group difference in accuracy for expected stimuli (post-hoc test of group effect for expected stimuli: *b*=0.239 (95%CI [0.04, 0.44], *z*=2.38, *p*=0.017; for unexpected stimuli: *b*=-0.038 (95%CI [-0.275, 0.199], *z*=-0.315, *p*=0.752). In other words, individuals with ASD performed as well as controls on unexpected trials when cue information was potentially misleading. However, they did not benefit as much from the cue during expected trials where the cue provided helpful information about the stimulus direction (Figure 4A).

**Figure 4.**
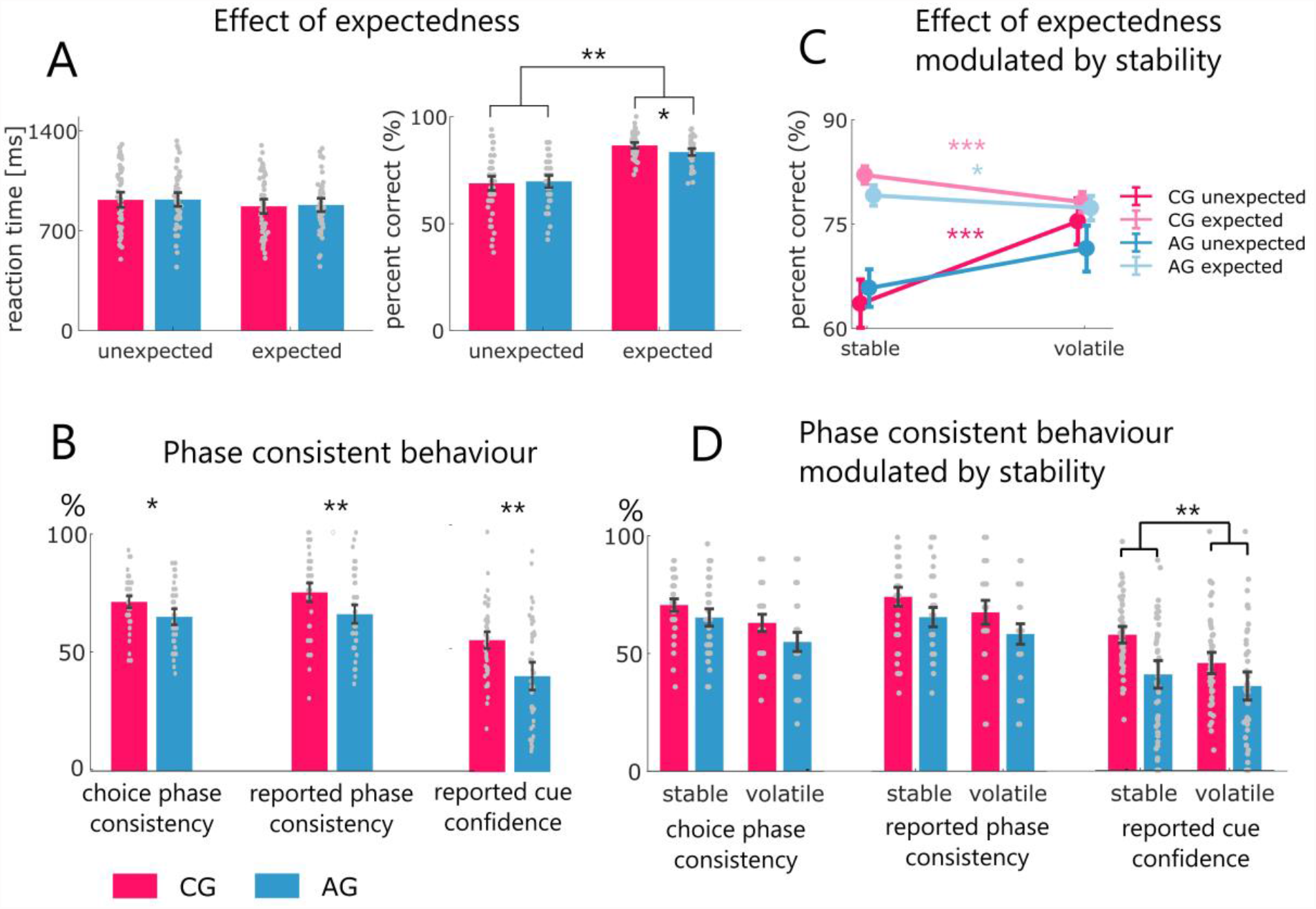
Group differences between expectedness and stability. Panel **(A)** shows the effect of expectedness on RT (left) and accuracy (right) (H2a). There is a significant interaction between expectedness and group on accuracy but not RT (see main text). Panel **(B)** shows phase consistent behaviour. “Phase consistent” means that the response is consistent to the phase of the cue (congruent in valid phases and incongruent in invalid phases) in trials without stimulus information, i.e. zero coherence trials (left) and questions (middle and right bar plot) (H2b, H2c). A more detailed description of phase consistency is given in the Methods section. Panel **(C)** depicts the significant three way interaction effect of expectedness x stability x group (*χ*^2^*(1)*=6.75, *p*=0.009) (H4a). Post-hoc tests revealed that volatility modulated the effect of expectedness more in the CG compared to the AG. Finally, panel **(D)** shows how the group effects of trials without stimulus information in panel B are modulated by stability (H4b, H4c). While there is no significant interaction effect of group and stability on choice phase consistency (direction response in zero coherence trials) and on reported phase consistency (direction response in question trials), we found a significant group x stability interaction effect of reported cue confidence, where stability modulated cue confidence more in the CG compared to the AG. Abbreviations: CG: control group, AG: autism spectrum disorder group.

This suggests that individuals with ASD take information by the cue less into account than controls.

### Individuals with ASD benefit less from cue information in the absence of sensory information about motion direction (H2c-e)

Until now, we have examined the effects of the use of prior information on performance in the presence of stimuli with non-zero motion coherence, where information provided by the cue competes with actual sensory information. Another more direct readout of the use of prior information are trials where sensory stimuli are not informative about motion direction, i.e. zero-coherence trials (H2c). We found that the AG were significantly less likely to base their decisions on the phase of cue validity (phase consistency on zero coherence trials: *t*_*96*_=2.52, *p*=0.013, *d*=0.512). This effect is mirrored by their subjective report during question trials (H2d; reported phase consistency: t-test: *t*_*96*_=2.77, *p*=0.007, *d*=0.562) and their reported cue confidence (H2e; reported cue confidence: Mann-Whitney U test: *U*=3.15, *p*=0.002, *r*=0.320) (see Figure 4B). This suggests that participants with ASD relied less on the prior information provided.

Importantly, this effect was not driven by difficulties of individuals with ASD in inferring the meaning of the cue during invalid phases, but rather by a general reduction in cue use (see additional post-hoc analysis in Supplementary Material: Reduced cue benefit not due to invalid phases only).

### Reduced reliance on cue is not a consequence of inflexible priors (H3)

Earlier work proposed that inflexibility in the use of prior information rather than general uncertainty might drive deficits in individuals with ASD.^11,19^ To test this, in the CMDT, the first experimental phase (49 trials) used a stably reliable cue (85%) to allow for the acquisition of an informed prior. We tested whether after exposure to this early phase the AG showed a tendency to choosing the cue-congruent motion direction more frequently over the rest of the experiment compared to the CG, independently of the validity of the phase. While there was a marginally significant effect, this was in the opposite direction as postulated by the hypothesis (H3a; *t*_*96*_=2.11, *p*=0.038, *d*=0.428), i.e. the CG tended to choose the cue congruent direction more often than the AG. Furthermore, and also not consistent with the hypothesis, there was no group difference in the cue consistency reported on question trials (H3b): *t*_*96*_=0.856, *p*=0.394, *d*=0.174). These findings suggest that the overall tendency to rely less on the cue in individuals with ASD was not driven by the inflexibility to change a prior established early in the task (as was predicted by hypothesis 3).

### Volatility-induced hierarchical learning is diminished in the AG (H4)

Finally, the last hypothesis suggested that overreliance on sensory information in individuals with ASD may be driven by alterations in hierarchical learning. To test this, our CMDT paradigm introduced volatile changes in cue validity. According to Bayesian models of learning, higher uncertainty about cue reliability should occur during volatile phases, accompanied by faster learning rates.

We first assessed the predicted effect of higher uncertainty about cue validity by comparing the expectancy effect on accuracy between stable and volatile phases (H4a). A GLMM (fixed effects: stability, expectedness, group; random effects: subject) showed that stability affected the expectancy effect (expectedness × stability interaction: *χ*^2^(1)=47.94, *p*=4.4e-12) and this effect was different across groups (3-way interaction: expectedness × stability × group: *χ*^2^(1)=6.75, *p*=0.009).

Interestingly, Tukey-corrected post-hoc tests showed that the 3-way interaction effect was driven by an opposing effect of stability on unexpected (*z*=4.70 *p*=1.5e-5) and expected trial outcomes in the controls (*z*=-5.41, *p*=3.7e-7) (Figure 4C): while overall performance for expected stimuli decreased during volatile phases, performance for unexpected stimuli increased. This suggests that during volatile phases the CG became less influenced by the cue. By contrast, in the AG, this effect of volatility was not observed for unexpected trial outcomes (*z*=1.96, *p*=0.204) and was less clear for expected trial outcomes than in CG (*z*=-2.80, *p*=0.026). This suggests that choices were less influenced by stability in the AG compared to the CG.

However, on trials without information about stimulus direction, i.e. trials with zero motion coherence (H4b) and question trials (H4c), a corresponding effect of stability on phase consistency was not found (Figure 4D; for further details, see Supplementary Material: Hierarchical learning hypothesis).

Finally, we examined reported cue confidence on question trials (H4d). We found a strong main effect of stability (GLMM: *F*_*1,95*_=33.93, 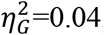, *p*=7.8e-8,) and group (*F*_*1,95*_=11.30, 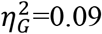, *p*=0.0011) and an interaction effect of group × stability (*F*_*1,95*_=5.94, 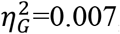, *p*=0.017) (Figure 4D).

Overall, hypothesis 4 could thus only be confirmed partially. While stability modulated accuracy and cue confidence differently in both groups, this effect was not apparent when examining phase consistency on trials without relevant sensory information.

## Discussion

In this study, we investigated competing predictions that arise from the general “Bayesian brain” theory of autism. While this theory suggests that autistic symptoms result from an overweighting of sensory information relative to prior information^8–10^, there are different reasons why this imbalance could arise. Specifically, four different causes have been proposed by previous literature: overprecise sensations (H1), imprecise priors (H2), inflexible priors (H3), and altered hierarchical learning (H4).

These hypotheses have been tested in previous studies,^19,22–25,27–29,31,32,43,44^ however, with conflicting results. At least the three aforementioned reasons may explain this inconsistency: (i) many studies did not compare the relative plausibility of the different hypotheses but tested a single one, (ii) some studies did not examine individuals with a clinical diagnosis of ASD, but autistic traits in the general population,^26,29,31^ and when diagnosed individuals were investigated, (iii) sample sizes were mostly small, ranging from *N*=14 to *N*=26 in most studies,^19,22,24,25,28,31–33,36,37,44^ with few exceptions.^23,27,34^

In the current study, we tried to address these issues and designed a set of experiments to systematically test the different hypotheses in a larger clinical sample (47 adults diagnosed with ASD and 50 matched controls). We found evidence for imprecise priors and, to a lesser degree, altered hierarchical learning, but not for overprecise sensations or inflexible priors. An overview of all hypotheses we tested and the results is provided by Table 3.

**Table 3.** Overview of the hypotheses tested in this study. The statements express the alternative hypothesis (in the context of frequentist null hypothesis testing). A green check mark indicates that the null hypothesis was rejected; a red cross indicates that the null hypothesis could not be rejected.

| **H1: Overprecise sensation hypothesis** | **H2: Imprecise prior hypothesis** | **H3: Inflexible prior hypothesis** | **H4: Hierarchical learning of priors** |
| --- | --- | --- | --- |
| *Does sensory processing and evidence accumulation differ between the AG and CG in the NCMDT?* | *Does the AG use contextual prior information less than the CGs?* | *Does the AG have more difficulties than the CG in updating a contextual prior after learning it initially?* | *Is the use of prior information in the AG less modulated by the volatility of the contextual cue than in the CG?* |
| ✗ **H1a**<br>The AG and CG differ in accuracy and RT.<br>✗ **H1b**<br>The AG and CG differ in DDM parameter estimates (drift, threshold). | ✓ **H2a**<br>The AG benefits less than the CG from prior information provided by cues.<br>✓ **H2b**<br>The AG shows a smaller difference in accuracy between expected and unexpected stimuli than the CG.<br>✓ **H2c**<br>Responses of the AG are less phase-consistent on zero-coherence trials than response of the CG.<br>✓ **H2d**<br>Responses of the AG are less phase-consistent on question trials than those of the CG.<br>✓ **H2e**<br>The AG reports lower cue confidence than the CG on question trials. | ✗ **H3a**<br>The AG chooses options consistent with the cue more frequently (are more cue consistent) than the CG.<br>✗ **H3b**<br>The AG is more cue consistent in their reports on question trials than the CG. | ✓ **H4a**<br>Stability modulates the accuracy difference between expected and unexpected trial outcomes less in the AG than in the CG.<br>✗ **H4b**<br>Stability modulates the phase consistency less in the AG than in the CG.<br>✗ **H4c**<br>Stability modulates the reported phase consistency less in the AG than in the CG.<br>✓ **H4d**<br>Stability of the phase modulates the reported cue confidence less in the AG than in the CG. |

In the following, we briefly discuss these results hypothesis by hypothesis and compare them to previous literature.

### Hypothesis 1: Sensory information processing is more precise in individuals with ASD

Sensory processing in individuals with ASD has been investigated by numerous studies.^45^ Although the literature is not consistent, several studies in different sensory domains suggest that individuals with ASD may experience hypersensitivity to sensations,^46–49^ may process low-level stimulus features with higher sensory precision,^15–17,46,50,51^ and may have difficulties in processing more complex sensory stimuli^50,52^ and integrating multiple sensory domains.^53,54^

Sensory processing can be investigated in many different ways. We chose a well-established experimental paradigm (moving dots task with different motion coherence levels^55^) that allowed us to quantify sensory processing using a drift diffusion model (DDM). Our analyses did not reveal any superiority of individuals with ASD in evidence accumulation – neither in overall accuracy and RT, nor in any of the DDM parameter estimates (H1a, H1b) – as would be expected when sensory precision is enhanced. On the contrary, there was a marginally significant effect (not surviving correction) in the opposite direction, i.e. the controls showed a faster accumulation of sensory information (higher drift). These results differ from those of a previous study using a smaller sample and a different paradigm (discrimination of orientation in static Gabor patches)^56^. Notably, this study focused on evidence accumulation as the only explanation of perceptual differences, without considering the possibility of altered priors. This is important because, from a Bayesian view, an increase in the weight of sensory information could equally result from enhanced sensory precision or less precise priors.

So far, only one study reporting enhanced sensory processing allowed for investigating changes in either sensory processing or priors.^26^ That study, however, used a different type of motion detection task and, more importantly, investigated autistic traits in the general population (as opposed to individuals with a clinical diagnosis of ASD).

### Hypothesis 2: Individuals with ASD weigh prior information less

The imprecise prior hypothesis assumes that perception in individuals with ASD is dominated by sensory information because their priors are less precise. This notion was first articulated, in an explicitly Bayesian context, by Pellicano and Burr.^10^ Their proposal was partially based on earlier studies suggesting that altered use of prior information may affect perception in ASD.^18,57,58^ Specifically, Pellicano and Burr^10^ proposed the existence of “hypo-priors” in individuals with ASD, i.e. priors with reduced precision, leading to smaller influence of prior expectations on perception and a greater weight of sensory information. They predicted that hypo-priors would become particularly visible in situations where priors help to resolve uncertainty. Our paradigm allowed for testing this hypothesis directly, confirming five distinct predictions (H2a-H2e):

First, comparing the versions of our task with and without cues, the AG benefitted less from the availability of cues than the CG (H2a), especially on trials with low coherence. Second, on trials where the outcome was predicted by the cue, the AG profited significantly less from the cue than the CG (H2b). Third, our experiment included trials where sensory input was completely uninformative (zero coherence trials) and the only information that could guide decisions was provided by the cue. On these trials, individuals with ASD relied significantly less on cue information than the controls (H2c). Fourth, on “question trials” in our experiment participants were asked, after the cue was presented, to indicate which stimulus direction they expected. In the AG, responses were less consistent with the cue prediction than in the CG (H2d). Fifth, on question trials, individuals with ASD reported lower confidence in their prediction than controls (H2e). Sixth, an additional model-based analysis (DDM with the starting value as free parameter to account for valid vs. invalid phases) provided results that were consistent with the model-agnostic analyses described above (see Supplementary Material).

Our results thus support the prediction by Pellicano and Burr^10^ that the precision of priors is reduced in ASD. This prediction has previously been tested by numerous studies using a variety of different visual or auditory tasks or temporal integration paradigms, both in individuals with a clinical diagnosis and in the general population. While the majority of studies yielded results consistent with less precise priors,^24,25,27,29,31–33,35^ two studies did not find evidence for weaker priors.^26,28^ We comment on possible reasons for this discrepancy below.

### Hypothesis 3: Individuals with ASD have inflexible priors

Inflexible priors might represent an alternative reason why prior information may not be exploited fully in individuals with ASD: they might be capable of acquiring priors upon exposure to a new environment or context, but are then possibly inflexible in adjusting their priors once this environment changes. This possibility was suggested by previous concepts and studies.^11,19,31,36^ Because inflexibility has been defined inconsistently in the context of Bayesian perception, the findings might also be explained by other causes,^36,37^ as discussed by some authors.^19^

We investigated the possible presence of inflexible priors by testing whether the priors acquired at the beginning of the experiment (during the initial valid cue phase) shaped responses throughout the remainder of the experiment. However, we did not find any evidence supporting this notion: individuals with ASD did not choose options consistent with the cue more frequently than controls, neither on standard trials nor on question trials (where the expected direction had to be indicated, in the absence of a stimulus).

### Hypothesis 4: Volatility-induced hierarchical learning is diminished in the AG

Another reason why individuals with ASD may differ in their use of prior information may relate to how they adapt to changes in uncertainty, specifically through learning hierarchical relations between environmental causes. For example, the updating of low-level priors (e.g. about concrete events in the world) depends on higher-order estimates of abstract quantities, such as environmental volatility.^59,60^ For ASD, it has been suggested that changes in learning occur at higher (i.e. more abstract) levels of perceptual hierarchies in particular, e.g. due to altered precision of beliefs about volatility.^14^

The results from our study were largely in line with this hypothesis. In particular, we found a group difference in how volatility affected cue use (i.e. a group×volatility×expectedness interaction): in the CG, the cue exerted less influence on responses during volatile phases, with decreased performance for expected stimuli and increased performance for unexpected stimuli. By contrast, this effect was significantly weaker in the AG – a finding that would be consistent with altered hierarchical learning in ASD and ensuing weaker weighting of prior expectations compared to sensory information, e.g. due to higher estimates of volatility.^8^

However, one would expect that this putative difference is also visible on trials without sensory stimulus information (question trials and zero coherence trials). Here, our results were mixed: while we did not find an effect of volatility on phase consistency, we did observe an effect of volatility on reported cue confidence. In summary, although not all predictions could be confirmed, no evidence to the contrary was found. Overall, our findings thus support the hypothesis of altered hierarchical learning in ASD.

In the previous literature, three studies have examined learning under volatile conditions in individuals with ASD, with contradictory results. Two studies on adults with ASD also found evidence for altered hierarchical learning.^22,34^ One computational modelling study demonstrated elevated learning of volatility and increased estimates of changes in volatility (meta-volatility) during an audio-visual learning task, suggesting that individuals with ASD learn to “expect the unexpected”.^22^ By contrast, a second study,^34^ reported that perception in individuals with ASD during an auditory serial discrimination was more strongly dominated by information from the past, implying reduced learning of volatility. Finally, a third study failed to find a difference in learning rate between volatile and stable phases during a visual associative learning task.^23^ This study differed from our and the other previous studies in that it investigated children (not adults) with ASD and did not use stimuli imbued with sensory uncertainty.

### Strengths and weaknesses of our study

Our study has strengths and weaknesses. One particular strength of our study is that it was designed to provide readouts allowing for both qualitative and quantitative tests of four competing hypotheses. In order to test the processing of sensory information, we focused on an established computational model (DDM) that was applied to data from an equally well-established perceptual paradigm (dot motion task). Conversely, to probe the role of priors, we included specifically designed trials for testing the influence of prior expectations in the absence of *informative* sensory information (zero coherence trials) and in the absence of *any* sensory information (question trials). These trials allowed us to probe putative effects of altered priors without the need of adopting a specific model of perceptual inference. This qualitative approach has the advantage of obtaining a directly informative readout and obviates the need of inverting a (potentially complex) model. A weakness of this approach to testing the influence of priors is that understanding the exact mechanism behind data in relation to hypotheses H2 (imprecise priors) and H4 (altered hierarchical learning) requires trial-by-trial modelling.

Additionally, we took care to ensure that our experimental assessments avoided potential confounds such as previously reported deficits in motor skills in ASD.^61–64^ Instead, we developed patient-friendly tasks that only required eye movements to express decisions.

Finally, it is worth emphasising that, compared to previous investigations, we recruited a large sample of adults with ASD, together with a control group carefully matched with regard to age, gender, and handedness. Diagnosing adults with ASD is challenging and requires particular expertise, and some of the variability in the literature may result from different diagnostic standards. In our study, diagnoses of ASD were carefully established and confirmed by a specialised psychiatrist.

### Summary and outlook

This study has compared different hypothetical mechanisms of altered perception in ASD all of which derive from the general “Bayesian brain” theory. Our findings suggest that perception in adults with ASD relies less on prior information and may adjust less to changes in the environment. These findings may help explain salient clinical features of ASD, such as higher intolerance of uncertainty^65–70^ and frequent feelings of being overwhelmed.^10^ Our study illustrates how competing explanations derived from the same theoretical (i.e. Bayesian brain) framework can be compared, using a combination of computational modelling and qualitative tests. If the present results can be confirmed in future studies of this sort, they might help to inform new treatment approaches, e.g. cognitive interventions specifically focused on developing more precise prior expectations.^8^

## Data Availability

The analysis code can be accessed on the GitLab repository of ETH Zurich (https://gitlab.ethz.ch/tnu/code/schneebelietal_biasd_mdt). Upon acceptance in a peer-reviewed journal, the anonymized data will be made available in the ETH Research Collection (https://www.research-collection.ethz.ch/), a freely accessible scientific repository that complies with the FAIR principles42.

## Abbreviations

2AFC: Two-alternative forced choice
AG: ASD group
AQ: Autism quotient
ASD: autism spectrum disorder
ASD-I: Autism spectrum disorder interview
BDT: block design task
BIASD: Bayesian inference in autism spectrum disorder project
CI: confidence interval
CG: control group
CMDT: cued motion discrimination task
CSI: cue-stimulus interval
d2-R: d2 revised (attention test)
DDM: drift diffusion model
DSST: digital symbol substitution test
ES: effect size
GLMM: generalized linear mixed effects model
ITI: inter-stimulus interval
NCMDT: non-cued motion discrimination task
RT: reaction time
WFPT: Wiener first passage time

## Acknowledgements

We thank Natalie Araya for her assistance with regulatory issues. Furthermore, we acknowledge financial support by the Stiftung Netzwerk Schizophrenie (HH), René and Susanne Braginsky Foundation (KES), and the University of Zurich (KES).

