## Supplementary Material for "Disentangling “Bayesian brain” theories of autism spectrum disorder"

*joint senior authorship

### Overview

Literature ........ **Error! Bookmark not defined.**

### Supplementary Methods

#### Study design

The data set analysed in this paper was part of a larger study on autism spectrum disorder (ASD). Overall participants had three appointments: a clinical interview and two experimental sessions. Before each experimental session, participants were asked to fill out a series of questionnaires using an online survey tool. We furthermore conducted a neuropsychological assessment after the experiment during the second experimental session (see Supplementary Methods: Neuropsychological tests). The two tasks analysed in this study were conducted sequentially during the second experimental session: first, the participants completed the non-cued motion discrimination task, followed by the cued motion discrimination task.

##### Inclusion/ Exclusion criteria

Inclusion criteria for both groups were: age between 18 and 65 years, written informed consent, and the ability to adhere to the study protocol. For the control group, specific inclusion criteria were: no current psychological disorder, no severe psychological disorder in the past, and no first degree relatives with autism spectrum disorder. Individuals with ASD were included if they had been diagnosed with ASD by an external clinician and had an IQ > 70. For each participant, the diagnosis was confirmed in a preceding interview session by a board-certified psychiatrist and specialist in ASD using the autism spectrum disorder interview (ASD-I[1]). Exclusion criteria comprised: regular (> 1 units per week) intake of drugs within the last three months (except alcohol and nicotine), present anti-dopaminergic medication, daily intake of alcohol more than three units per day on average (1 unit=3dl beer=1dl wine), severe past or present neurological disorder, brain injury or neurological surgeries, and severe acute or chronic disease.

Out of the 111 participants, thirteen had to be excluded from the analysis: Three participants could not perform the task because their glasses were incompatible with the eye tracking system (to respond during the task, participants were required to do a saccade to the left or the right). Four participants were excluded due to other technical issues with the eye tracker. For one participant the data of the motion discrimination tasks were not properly saved. One participant was excluded because there were too many missing trials (no response in the predefined response time window in more than 10% of the trials). Lastly, as pre-specified in the analysis plan, to assure that all included participants performed and attended to the task, we excluded participants with too many incorrect trials (<65% correct trials) (n=2) and participants that did not reach a pre-set performance level in the simplest task conditions (n=2; minimal performance >60% correct responses for the random dot stimuli with highest coherence level, see Methods section).

#### Hypotheses testing

This section was adopted verbatim from the newest version of the analysis plan (see Supplementary Methods: Analysis plans) and reproduced here for completeness.

H1: First, we tested the *overprecise sensation hypothesis.* A corollary of this hypothesis is that sensory processing itself would be more accurate in individuals with ASD, independent of alterations in prior knowledge. We thus expected that indicators of evidence accumulation during the non-cued motion discrimination task (NCMDT) would differ between the two groups. Specifically, that individuals with ASD should have a higher accuracy and accordingly shorter reaction times (H1a), which should also be reflected in a higher drift rate in the Wiener first passage time (WFPT) (H1b).

H2: The *imprecise priors hypothesis,* contrarily proposes that an emphasis on sensory information is not caused by an increased sensory precision, but due to a decreased precision associated with prior knowledge, also referred to as ‘weak' or ‘flat’ prior.[2] A weak prior would therefore suggest that even when prior information is available, e.g. in form of the cue, it will be used less. Here this would result in a decreased benefit from the cue in the cued motion discrimination task (CMDT). One way to test this is to compare performance between the NCMDT and CMDT task, especially in low coherence trials, where the cue provides useful prior knowledge about the motion direction (H2a). Furthermore, less use of the cue should also reveal in a comparison between expected and unexpected trials. Specifically, if the prior information provided by the cue is used less by individuals with ASD the performance benefit (and thus difference) between expected and unexpected trials should also be smaller (H2b). Another indicator for this hypothesis could be observed in trials where direct motion evidence from the stimulus is absent (zero coherence stimuli and confidence questions trials), and the cue is the only indicator of motion direction. If individuals with ASD struggle using prior knowledge from the cue their responses should be less consistent with the cue during zero coherence trials (H2c) and during confidence questions (H2d) and their subjective confidence should be lower (H2e).

H3: According to the *inflexible prior theory,* individuals with ASD would learn a prior initially but have problems updating it over time. Here, this would mean that individuals with ASD would learn the first valid phase of the cue but have trouble updating this later during invalid and volatile phases. This would reflect in more cue consistent behaviour compared to the control group (CG) across the entire paradigm, especially at later phases (H3a) and in more cue consistent responses during question trials (H3b).

H4: While the differences mentioned in H2 could be an indicator of flat priors, they might also support the *hierarchical learning hypothesis*, where individuals had difficulties adjusting their priors according to difficulties in hierarchical learning. To disentangle the two we introduced volatility of the cue validity, using stable and volatile phases during the CMDT. Stability is an environmental factor, which is thought to modulate the speed of learning.[3,4] If the world is stable, priors become more precise and the learning rate, which can be expressed by the sensory precision divided by the prior precision, becomes smaller. In a volatile environment, priors become more uncertain and therefore, the learning rate is higher. If this modulation is affected as proposed by H4, the difference in accuracy and reaction time between expected and unexpected stimuli in volatile and stable phases would be smaller (H4a). We should observe a similar effect for phase consistency in trials without relevant sensory information, i.e. the modulation of phase consistency by stability should be smaller if hierarchical modulation of priors is weaker (H4b). This would also apply to the reported phase consistency in the intermitted question trials (H4c). Finally, the hierarchical learning hypothesis predicts that stability modulates certainty (measured by reported cue confidence) in individuals with ASD less compared to non-affected controls (H4d).

Lastly, if there are two processes contributing to ASD such as over-precise sensory input and imprecise prior, we should see effects in both, processing of sensory information and in at least one of the hypotheses predicted by the aspects of the imprecise prior theory.

#### Neuropsychological tests

We used four neuropsychological tests to exclude that differences in cognitive and intellectual abilities between the two groups were influencing the results in this study. Specifically, we tested perceptual logical thinking (BDT; block design task), working memory (arithmetic task), processing speed (DSST; digital symbol substitution test) from the HAWIE-R[5] and attentive and concentration skills (d2-R).[6]

In the DSST participants were required to transfer symbols to associated numbers as fast and accurately as possible using a pencil. In the BDT, participants were required to copy patterns using dices. Therefore, they had to mentally split the overall pattern into its local components. The arithmetic scale uses orally presented maths problems presented in a text format. To solve the task, participants must be able to comprehend the task, identify the question, keep the numbers in working memory and solve the arithmetic problem. Finally, we used the d2-R to measure attentive and concentration skills. In this paper-pen task, we asked the participants to sequentially find all $d$'s with two markings within $p$'s and $d$'s with 1-3 markings. The task requires attention, accuracy, speed and concentration. For group comparison, we acquired the overall measure F which is calculated as: $F = \frac{N_{missed}+N_{error}}{N_{targets}}$.

The two groups were balanced according to the measured intellectual abilities (see Table 1), with the exception of the DSST. Besides speed processing, this task requires fine motor skills. We presume that the observed group difference may reflect a difference in fine motor skills, as they have been shown for autistic patients,[7,8] rather than intellectual ability.

#### Apparatus

Participants' responses during the NCMDT and CMDT were measured by eye saccades towards the left or right side. This response type (as opposed to motor responses) was chosen because of frequently reported motor problems in individuals with ASD.[8–10] Experimentation took place in a dimly illuminated room. Participants viewed stimuli on a CRT screen (41.4x30cm; Philips 20B40, 85Hz) with 60cm distance. They were recorded with an infrared eye tracker (Eyelink 1000, SR Research, Ottawa, Canada) with a sampling rate of 1000 Hz. A chin rest was used to stabilize head position with 60cm distance from the screen. Both tasks and the training trials started with the 5-points calibration procedure from the Eyelink system (User Manual; SR Research, 2010). The experiment was programmed in MATLAB version R2016b and Psychtoolbox-3.

#### Task design

##### Non-cued motion discrimination task (NCMDT)

On every trial of the NCMDT (Figure 2A), participants were asked to fixate on a small point at the centre of the screen (0.7deg visual angle) between two Pac-Man like shaped target locations (11.3 degrees displaced from the centre on the left and right side with a radius of 3.5

degrees) (see Figure 2). After a random fixation period between 300-1200ms (ITI, drawn form a uniform distribution), a random dot motion stimulus was presented in a virtual aperture in the centre of the screen (7 degrees). Notably, if the participant did not fixate the dot at the end of the ITI (as detected by eye tracking), additional fixation periods of 100ms each were added until the participant successfully fixated the dot. The average movement direction of the dots could be leftward, rightward or no direction and was created by moving a coherence-dependent subset of white 2-pixel sized dots in each video frame coherently towards one direction. The dot density was 16.7 dots/deg2/s and the displacement of the coherently moving dots produced an apparent speed of 5deg/s. The stimulus presentation was implemented using the Shadlen toolbox (Shadlen Lab at Columbia University, New York, https://shadlenlab.columbia.edu/resources/VCRDM) and has been described in detail in previous studies (Resulaj, Kiani, Wolpert, & Shadlen, 2009; Roitman & Shadlen, 2002).

We used seven stimulus difficulty levels corresponding to different motion coherence levels in percent ([-51.2, -12.6, -3.2, ±0, +3.2, +12.6, +51.2]%), which occurred equally often across the whole experiment. Positive numbers denote an average motion direction to the left, negative numbers an average motion direction to the right (Figure 2B). Participants were instructed to indicate the motion direction via a saccade to the left or right target location as soon as they made their decision. Participants were instructed to respond as accurately and fast as possible. Immediately after a saccade to one of the target locations was made, the Pac-Man symbol in that location would close its mouth to signal that a decision had been detected. Once participants looked back to the fixation point at the centre of the screen, they were provided with trial-by-trial feedback after a 300ms fixation period: thumbs up for a correct response or thumbs down for an incorrect response, centred on the screen. The feedback for the zero coherence stimulus was randomly selected to be positive or negative. If participants failed to respond within a 2500ms response period, the stimulus presentation was stopped, a “too slow” message was shown at the centre of the screen, and the trial was stored as a time-out trial. Feedback, (positive, negative and too slow trials) was shown within a circle with a radius of 2.1deg.

Participants observed a progress bar of their performance at the top of the screen throughout the task: correct responses led to a reward of 10 points, whereas incorrect responses or timeouts resulted in a loss of 10 points. For each completed progress bar (100 points), participants received a bonus of 0.10 Swiss Francs in monetary reward that was paid out at the end of the experiment. Participants first played 280 trials of the non-cued version of the motion discrimination task, including 40 repetitions of each motion coherence level.

**Cued motion discrimination task (CMDT)**

The cued motion discrimination task was identical to the non-cued version of the task, except that after the fixation interval and prior to each stimulus presentation an arrow cue (2.1 degrees) appeared for 400ms at the centre of the screen. To ensure that participants fixated the centre of the screen when the cue appeared, the fixation period was repeatedly prolonged by 100ms if the participant did not fixate on the cue within 100ms at the end of the 400ms block. The cue was followed by a cue-stimulus-interval (CSI) of 1000ms (Figure 2C). The arrow cue pointed towards the left or the right and provided additional information about the upcoming motion stimulus. The validity of the cue (probability of pointing towards the correct motion direction) was varied over the course of the game: It started with a stable valid phase (49 trials, 85% cue validity), followed by a stable invalid phase (49 trials, 15% cue validity), followed by a volatile phase of alternation between valid and invalid phases (3 repetitions with 14 trials each), followed by two more periods, one stable valid (49 trials, 85% cue validity) and one stable invalid (49 trials, 15% cue validity). This resulted in 9 phases and 266 trials in total (see Figure 2D & E). Importantly, feedback of zero coherence trials was adjusted to the phase. For instance, during an invalid phase (cue validity=15%), a left response after a leftward cue in a zero coherence trial would be rewarded in 15% of the cases. The overall feedback (leftward or rightward motion correct) for zero coherence trials, was balanced throughout the experiment.

In 15 trials in total (evenly distributed across the full trace), instead of the random dots stimulus, a question probing the participant's confidence in predicting trial outcome was presented right after the cue presentation and CSI (see Figure 2F). The following text was presented: "In which direction will the dots move next? Place the point further out the more confident you are." Participants responded using the arrow keys on the keyboard to move a cursor along a continuous line (13.3 degrees) to the left or to the right (starting location of the cursor was always at the centre of the bar, corresponding to no motion direction and maximal uncertainty; see Figure 2D). The dot could be placed on any point along the line. Minimal step size of the dot was 1% of the line (e.g. 0.13 degrees). To move the dot from the centre to the left or right maximum, the arrow button had to be pressed for approximately 1500ms. Moving the dot back and forth was allowed. Participants indicated by button press as soon as they were satisfied with their choice. The overall position (left or right) indicated their expected motion direction, while the distance from centre (e.g., far out) indicated the confidence (e.g., high for far out). In a training phase prior to the task, participants were instructed how they could indicate both motion direction and confidence on this scale.

#### Drift Diffusion Model

We fit the data using a Drift Diffusion-Model.[12,13] The key idea behind this model is that in a decision process, $x(t)$, evidence is accumulated according to a Wiener diffusion process with a drift $v$ and a starting point $z_0$. This process ends if a threshold value $b$ is reached (i.e. as soon $x(t) > b$ or $x(t) < 0$). If the variance of the Wiener diffusion process $\sigma^2$ is fixed to 1 and the starting point $z$ at $x(0)$ lies between 0 and $b$, this process is referred to as Wiener first passage time[14] (WFPT). Whereas the calculation of the distribution of reaction times with the full drift diffusion model requires a large number of trials,[15] a simpler and faster method was provided by Navarro & Fuss[14] to calculate the probability density function of reaction times with the WFPT. Because the original analytical expression of the WFPT distribution $f(t|v, b, z)$ provided by Feller[16] contains an infinite sum (and is therefore not computable in finite time), Navarro & Fuss[14] provide an approximation, which is implemented in two different functions: one function is optimized for longer and one is optimized for shorter reaction times. This method was translated into different programming languages such as MATLAB, R and Python[17,18] and used in experimental settings.[19,20] Figure S 1 shows an example of the Wiener first passage time model.

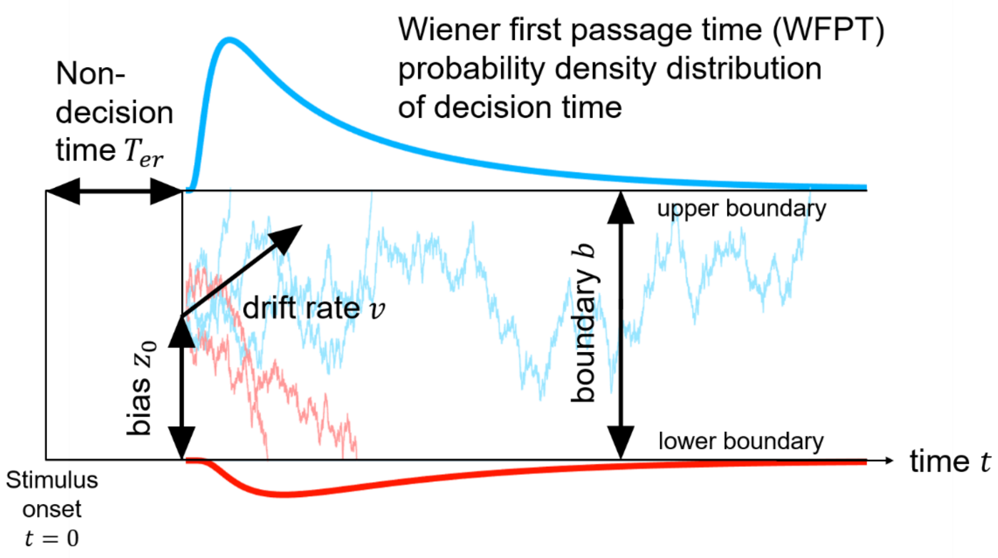

Figure S 1 A schematic of the Wiener first passage time distribution (WFPT). After a certain non-decision time ($T_{er}$) the Wiener diffusion process, i.e. the evolution of state $x(t)$ (in blue/red) starts. The starting point $z_0$ represents the bias where the process starts at time $x(0)$. A value higher than $\frac{b}{2}$ means a bias towards the upper boundary, and a value lower than $\frac{b}{2}$ is a bias towards the lower boundary. If $x(t)$ surpasses the value of the boundary $b$ or falls below zero, the decision is made. The drift $v$ represents the speed of evidence accumulation. A higher drift means faster accumulation.

To fit the parameters of the WFPT we used a log-likelihood-function procedure using the genetic algorithm (*ga*) of the MATLAB optimization toolbox. We minimize the sum of negative log likelihoods over trials $t$:

$$f(rt, y, b, v, z_0, T_{er}) = -\sum_{t=1}^{n} \log(y^{(t)} \cdot wfpt_{up}^{(t)} + (1 - y^{(t)}) \cdot wfpt_{low}^{(t)}) \quad \text{Eq. S 1}$$

where the superscript $t$ is the trial index, $wfpt_{up}$ is a function of reaction time $rt^{(t)}$, boundary b, drift rate $v$, starting point $z_0$ and non-decision time $T_{er}$. $wfpt_{up}^{(t)}$ denotes the probability of the reaction time $rt^{(t)}$ if the upper boundary is hit (if $y = 1$) and $wfpt_{low}$ if the lower boundary is hit. Importantly, the starting point z was fixed to $\frac{b}{2}$, assuming no initial directional bias.

We fitted the DDM without starting point to both tasks (cued and uncued) to assess performance differences across tasks (see Supplementary Material: Results). Furthermore, we fitted the DDM with z as free parameter. The starting point was then calculated as:

$$z_0 = \begin{cases} (1 - z) \cdot a & phase = 0\ (valid) \\ z \cdot a & phase = 1\ (invalid) \end{cases}$$

Eq. S 2

where $z$ is the free parameter and defines the strength of the bias, a is the boundary and $z_0$ is the effective starting point of the diffusion process. Consequently, the higher the estimate of parameter $z$, the more the underlying structure of the cue is learned.

#### Parameter recovery analysis

We validated the two models used in this work – WFPT with bias and without bias (i.e. the bias was fixed to 0.5) – according to the guidelines of Wilson and Collins[21]. One hundred data sets were simulated by drawing one sample per trial from the WFPT distribution (280 trials for the NCMDT and 266 trials for the CMDT) for each data set. For the 100 simulations, we used the parameter range from the actual participant's sample. Then we used the same maximum likelihood approach to recover the parameters as we used to fit the models (in-built optimization algorithm from MATLAB *ga*). We did this procedure for the simple WFPT model with fixed starting point (e.g. boundary b, drift v and non-decision time $T_{er}$ as free parameters) and for the WFPT model, where the starting point represented a bias towards the current phase (see also main text). Model recovery was successful, simulated and recovered parameters correlated between *r* = 0.97 and *r* = 0.99.

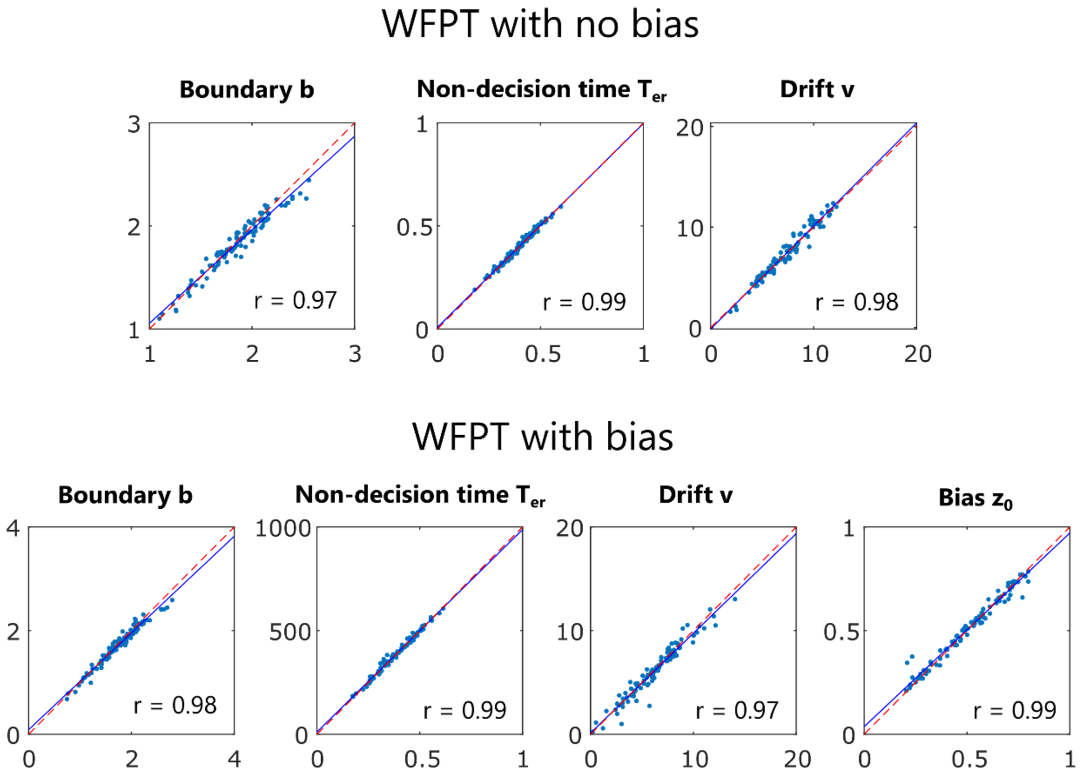

**Figure S 2 Parameter recovery.** Correlation between simulated and recovered parameters (boundary b, non-decision time $T_{er}$ and drift v) for the simple WFPT model with fixed starting point and for the WFPT model, where the starting point reflected a bias towards the current phase over the cued task (i.e. valid bias in valid phases and inverted bias in invalid phases).

#### Outlier Removal

Prior to the analysis, and as pre-specified in the analysis plan, we removed outliers for each participant (2.5% shortest reaction times and 2.5% longest reaction times per participant) in order to only keep the 95% interquantile.[18] Additionally, missing trials (i.e. timeouts) were also excluded from the analysis. On average, we excluded 6.34 missing trials (mean: 2.3%, std: 2.6%) in the NCMDT and 3.14 trials (mean: 1.2%, standard deviation: 1.7%) in the CMDT. There was no significant difference in the number of excluded trials across the two groups (Wilcoxon rank-sum test: NCMDT: $U$=1.25, $p$=0.211; CMDT: $U$=1.45, $p$=0.146).

#### Analysis plan

The original time-stamped analysis plan (Analysisplan_SUT.docx, https://gitlab.ethz.ch/tnu/analysis-plans/schneebelietal_biasd_mdt; clean version:

Analysisplan_SUT_v1.docx) was more extensive than the results reported here. The tests which were not performed and the exact reasons for revisions are referred to in detail in the second and third version of the analysis plan (Analysisplan_SUT_v2.docx). Importantly, when writing the paper, we changed the sequence and the labelling of the hypotheses for better structure and understanding. To avoid confusion, a third analysis plan was uploaded that contains the final structure and numbering of hypotheses (Analysisplan_SUT_v3.docx). Details on the exact updates are given in the revision section of the analysis plan.

### Supplementary Results

#### Performance difference across tasks

First, we tested if the task manipulations were successful across the whole sample. We therefore first calculated a paired t-test between the cued and non-cued task with percent correct (PC) and reaction time (RT) as dependent variables. Note that for the following analyses zero-coherence trials were excluded for PC but not RT as dependent variable, because correctness was randomly assigned for these trials. We found an increase in performance ($t_{96} = -2.36, p = 0.020, d = -0.25$; $\overline{PC}_{non-cued} =$ 81.1 (± 7.4)%; $\overline{PC}_{cued}$ = 82.8(± 6.2)%) and a decrease in reaction time ($t_{96} = -2.36, p = 0.020, d = -0.25$; $\overline{RT}_{non-cued} = 1040\ (\pm\ 192) ms$; $\overline{RT}_{cued} = 914\ (\pm\ 213) ms$). This suggests that on average the information provided by the cue was used and helped increase speed and accuracy in motion discrimination. Next, we included motion coherence and calculated a linear mixed effect model for RT and a generalized linear mixed effects model for PC with task and coherence level as fixed factors and subject as random factor. There was a strong main effect of coherence on RT (Greenhouse Geisser corrected ($\varepsilon$=0.24, $p$<2.2e-16) main effect of coherence: $F_{1.71,164.4}$ = 353.0, $p$=4.32e-56, $\eta^2_G$=0.44) and PC (main effect of coherence: $\chi^2(2)$=6926.2, $p$<2.2e-16). Importantly, there was an interaction effect of task and stimulus difficulty level on RT (mixed model: Greenhouse-Geissner corrected ($\varepsilon$=0.75, $p$=7.7e-13) interaction effect: $F_{2.3,217}$=28.75, $\eta^2_G$=0.02, $p$=2.7e-16) and PC ($\chi^2(2)$=21.29, $p$=2.38e-5, Table S1).

Post-hoc test analysis revealed that this interaction effect was particularly driven by stimulus levels with high difficulty (low motion coherence). In RT, there is an effect for all but the highest coherence level (0% coherence: $t_{197.4}$=9.81, $p$=9.4e-19; 3.2% coherence: $t_{197.3}$=8.27, $p$=2.0e-14; 12.8% coherence: $t_{197.3}$=5.30, $p$=3.1e-7; 51.2% coherence: $t_{197.3}$=1.76, $p$=0.080) (**Figure S 3**). In PC, there was a strong effect for the most difficult level (3.2% coherence: $z$=-6.55, $p$=5.7e-11) but not for the levels with lower difficulty (12.8% coherence: $z$=-0.13, $p$=0.900; 51.2% coherence: $z$=1.53, $p$=0.13). This suggests that the increase in performance caused by the additional cue information could be observed especially on trials with low coherence.

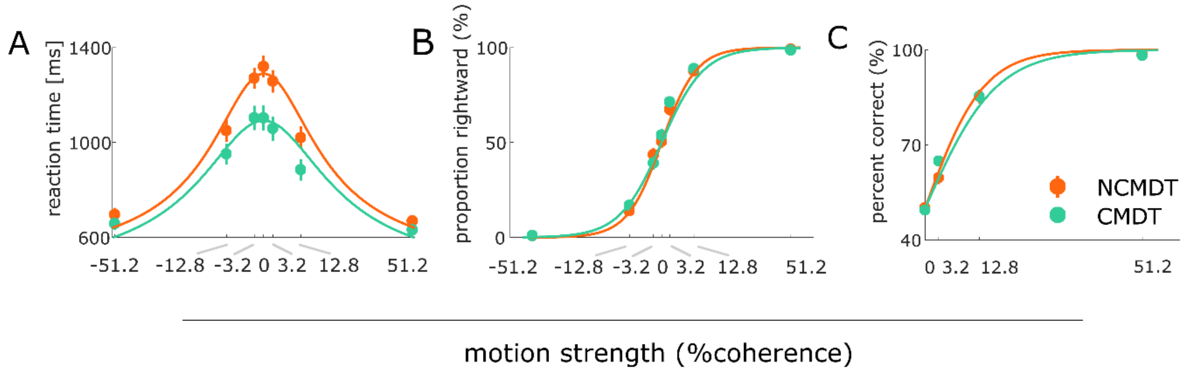

**Figure S 3 Collapsed data of all subjects during the non-cued (orange) and cued (green) version of the motion discrimination task**. Solid line indicates fit by the WFPT: (A) Reaction time across motion coherence levels; (B) Proportion rightward responses across motion coherence; (C) Proportion correct responses across unique motion coherence (left and right combined). Chance performance was assigned to zero coherence stimuli. Error bars reflect 95% confidence interval.

This effect was also reflected in the model-based analysis. Across the two tasks, we found a significant difference with regard to the boundary parameter ($t_{194}$=3.54, $p$=0.00049, Cohen's $d$=0.506), but not the drift rate of the DDM ($t_{194}$=1.49, $p$=0.137, Cohen's $d$=0.213). Furthermore, the non-decision time was significantly different between both tasks ($t_{194}$=2.21, $p$=0.028, Cohen's $d$=0.32).

| | NCMDT | CMDT | $T_{194}$ | $p$ | $d$ |
|---|---|---|---|---|---|
| boundary $b$ | 1.89 (0.32) | 1.70 (0.39) | 3.54 | 0.00049 | 0.506 |
| non-decision time $T_{er}$ [ms] | 392.9 (8.7) | 367.2 (7.5) | 2.21 | 0.028 | 0.32 |
| drift $v$ | 7.29 (2.50) | 6.78 (2.26) | 1.49 | 0.137 | 0.213 |

**Table S 1 Summary statistics of the WFPT with fixed starting point**. We used paired t-tests for all comparisons. NCMDT: Non-cued motion discrimination task; CMDT: cued motion discrimination task

#### Overall effect of validity, stability and expectancy in the CMDT

To alter the content and reliability of the a-priori information the cue in the task was not stable but changed its validity. The cue was correct in 85% during a valid phase and 15% correct during an invalid phase (Figure S 3D). Moreover, these probabilities changed in time such that there were stable and volatile phases in which the cue validity altered slowly or rapidly. These manipulations served to detect how much an individual would rely on the cue ignoring its validity (cue consistency), how much they would be able to use the cue taking its validity into account (phase consistency; outcome expectancy), and to what extent they would alter their cue use during stable and volatile episodes.

**Validity**

Overall participants successfully learned whether the cue was in a valid of invalid phase. When sensory uncertainty was high (during zero coherence trials), motion direction responses were significantly more likely to follow the recommendation of the cue during valid (70.9(±15.3)%) than invalid (40.8(±14.9)%) phases (t-test: $t_{96}$=12.31, $p$<2e-16, d=1.99). Similarly, when asked about the expected motion direction during question trials the self-reported belief reflected the estimated cue validity (reported phase consistency (valid): 74.11(±19.3)%; reported phase consistency (invalid): 38.0(22.9)%; $t_{96}$=10.81, $p$<2e-16, $d$=1.71). This suggested that participants detected and reacted to changes in the validity of the cue while they still showed an overall bias to follow the cue in particular in the absence of sensory information.

**Expectancy**

To test expectancy of the stimulus, we compared trials where the prediction of the cue was surprising (valid cue during an invalid phase or invalid cue during a valid phase). Comparing expected versus unexpected trial outcomes, accuracy was significantly higher (Wilcoxon signed rank test, $U$=8.05, $p$<8.3e-16, $ES$=0.58) and reaction times were significantly lower (paired t-test, $t_{96}$=5.60, $p$<9.2e-8, $ES$=0.14). To ensure, that we did not accidentally remove unexpected trials or that results were distorted because of too many missing trials in the unexpected condition, we compared the number of missing trials and number of outliers between these conditions. Both showed no significant differences (missing trials: Wilcoxon signed rank test, $U$=1.29, $p$=0.196, $ES$=0.09; outliers: $U$=1.34, $p$=0.180, $ES$=0.10).

**Stability**

Furthermore, we tested the manipulation of stability. In this case, we tested the difference of phase consistency, i.e. the tendency to choose the direction of the current phase in trials without sensory information and reported certainty between stable and volatile phases. Individuals were more uncertain in the volatile than in the stable phase in their reports (t-test: $t_{96}$=5.76, $p$=1.0e-7, $d$=0.39) and were less consistent to the phase in zero coherence trials (t-test: $t_{96}$=5.22, $p$=1.0e-6, $d$=0.60) and question reports (t-test: $t_{96}$=3.71, $p$=0.00035, $d$=0.37).

**Relative time point**

The relative time point was defined as the 25 trials (half of the stable phase) before and after the volatile phase. It has been shown that uncertainty, and therefore also the learning rate, is increased after a volatile phase in Bayesian learners.[3,4] The manipulation of the relative time point was partly successful. It was significant for cue confidence (Wilcoxon signed rank test: *U*=-2.50, *p*=0.0125), but not for reported phase consistency (Wilcoxon signed rank test: *U*=-0.539, *p*=0.590) nor choice phase consistency (t-test: $t_{96}$=1.90, *p*=0.060, Cohen's *d*=0.23). Note that within the 25 trials before and after the volatile phase, only four trials were zero coherence trials and only two trials were question trials. Because of this limited number of trials per condition, these results must be considered with caution.

#### Reduced cue benefit in the ASD group is not driven by invalid phases only

To test if the effects for hypothesis H2b and H2c in the main paper are driven by difficulties in inferring the meaning of the cue in invalid phases, we used a generalized mixed model with an additional factor for cue validity. We found evidence for an overall decrease of prior use independent of cue validity in the autism group (AG; three-way interaction: expectedness × cue validity × group: ($\chi^2(1)$ =1.40, *p*=0.24). In line with the results reported in the main text, we found a significant interaction between expectedness and group ($\chi^2(1)$=11.26, *p*=0.0008). Interestingly, there was also a strong interaction effect of expectedness and validity of the phase ($\chi^2(1)$=43.15, *p*=5.1e-11). Although a bias towards the current phase was present in both invalid and valid phases, the effect seemed to be stronger for valid (*b*=-1.31, (95%CI [-1.44, -1.19], *OR*=0.27, *p*<0.0001) than for invalid phases (*b*=-0.70, (95%CI [-0.83, -0.58]), *OR*=0.49, *p*<0.0001). Furthermore, there was an interaction effect of group and phase validity ($\chi^2(1)$=7.50, p=0.006). This interaction effect was driven by a higher benefit from the prior in valid phases than invalid phases in the CG compared to the AG (post-hoc test AG vs. CG in expected, valid trials: *b*=0.46, (95%CI [0.23, 0.68]), *OR*=1.58, *p*=0.0001). In all remaining categories (expected and unexpected trials in invalid phases, and unexpected trials in valid phases) no significant group differences were found.

To test whether the increased cue use of the CG in valid phases was consistent on trials in the absence of sensory information, we furthermore investigated if phase consistency was modulated by phase validity in zero coherence trials and question trials. We did not find any interaction between phase validity and group for phase consistency during zero coherence trials ($\chi^2(1)$=2.20, *p*=0.14) nor question trials ($\chi^2(1)$=0.79, *p*=0.37). There was also no interaction effect of phase validity and group on cue confidence on question trials ($\chi^2(1)$=0.10, *p*=0.75).

In summary, by introducing phase validity into the model, we confirmed that the group effect on expectedness was not due to a problem in inferring the meaning of the cue during invalid phases in individuals with ASD; instead they relied less on prior information.

#### Phase consistency is not mediated differently by volatility between groups

The hierarchical learning hypothesis predicts that the AG is less able to update their learning strategy in volatile phases compared to the CG. This hypothesis was partly confirmed. While there was a three-way interaction effect of expectedness × stability × group, confirming H4a, (see main text), in trials without relevant sensory information about the direction, i.e. in zero coherence trials (H4b) and question trials (H4c), we did not find the corresponding interaction effect. In a generalized linear mixed effects model (GLMM; with fixed factors: stability and group; random factor: subject) a group × stability interaction was not found in either model (H3b; phase consistency: $\chi^2(1)=0.40$, $p=0.53$; H4c; reported phase consistency: $F_{1,95}=0.02$, $\eta_G^2<0.0001$, $p=0.88$). However, there was a main effect of stability ($\chi^2(1)=25.89$, $p=3.6e\text{-}7$) and main effect of group ($\chi^2(1)=6.43$, $p=0.011$) on phase consistency. Corresponding main effects were found for reported phase consistency (main effect of stability: $F_{1,95}=13.64$, $\eta_G^2=0.03$, $p=0.0004$; main effect of group: $F_{1,95}=7.63$, $\eta_G^2=0.06$, $p=0.007$. In other words, the changing validities of phases were less recognized in the volatile phases across both groups, while the AG had more difficulties to recognize phase validities independent of phase stability, but there was no unique effect of the AG on stability.

#### No group effect of the relative time point

We calculated mixed effects models with choice phase consistency in zero coherence trials, reported phase consistency and cue confidence in question trials as dependent variables. Fixed effects were the trials before and after the volatile phase (relative time point) and group, while subject was a random effect. Note that per condition there were only very few trials (four and two, respectively) and the results must be interpreted with caution.

There was no interaction effect of group and relative time point on choice phase consistency (generalized mixed model: $\chi^2(1)=0.05$, $p=0.83$) nor on reported cue confidence (mixed model: $F_{1,95}=0.82$, $p=0.37$, sphericity assumed). Finally, there was a near-significant interaction effect on group of the relative time point and group on reported phase consistency (generalized mixed effects model: $\chi^2(1)=3.35$, $p=0.07$).

#### Learning as starting point: Smaller bias towards the current phase in the AG

Learning across trials would be reflected as the change in the starting point of the WFPT model. To assess learning over the whole task, we introduced a starting point parameter $z_0$ (WFPT model with free starting point). This parameter reflects the bias towards the expected stimulus, i.e. the bias towards the cue direction in valid phases and the bias against the cue direction in invalid phases. A value of 0.5 would indicate no bias and ignorance towards the validity of the cue. A value larger than 0.5 and smaller than 1 would indicate a bias towards expected trials. In the hypothetical case where the stimulus is ignored completely and the decision to go with the expected direction has been made in advance, $z_0$ would result in a value of 1.

The starting point model parameter $z_0$ captures the bias towards expected trials in both groups but also the group differences, i.e. the CG developed a slightly stronger bias over the whole experiment (t-test: $t_{96}$=2.34 $p$=0.021, Cohen's $d$=0.48). These modelling results reflect the behavioural results and contribute to a better understanding of the processes of evidence accumulation in the both groups.

| | | All (*n*=97) | CG (*n*=50) | AG (*n*=47) | $t_{96}$ | *p* | *d* |
|---|---|---|---|---|---|---|---|
| NCMDT | *b* | 1.88 (0.32) | 1.88 (0.34) | 1.89 (0.29) | 0. 17 | 0.87 | 0.03 |
| (no bias) | *v* | 7.29 (2.50) | 7.79 (2.22) | 6.77 (2.69) | 2.04 | 0.04 | 0.41 |
| | $T_{er}$ *[ms]* | 393 (87.5) | 409 (63.6) | 376 (105.0) | 1.89 | 0.06 | 0.38 |
| CMDT | *b* | 1.70 (0.39) | 1.70 (0.42) | 1.70 (0.36) | 0.003 | 1.00 | 0.001 |
| (no bias) | *v* | 6.78 (2.26) | 7.06 (2.19) | 6.50 (2.31) | 1.25 | 0.21 | 0.25 |
| | $T_{er}$ *[ms]* | 367 (75.1) | 368 (67.3) | 366 (83.2) | 0.11 | 0.92 | 0.02 |
| CMDT | *b* | 1.74 (0.39) | 1.74 (0.41) | 1.74 (0.37) | 0.005 | 1.00 | 0.001 |
| (with bias) | *v* | 6.50 (2.28) | 6.76 (2.22) | 6.24 (2.32) | 1.14 | 0.26 | 0.23 |
| | $T_{er}$ *[ms]* | 370 (82.9) | 376 (67.6) | 363 (96.7) | 0.79 | 0.43 | 0.16 |
| | $z_0$ | 0.55 (0.04) | 0.56 (0.04) | 0.54 (0.03) | 2.34 | 0.021 | 0.48 |

Table S 2 Model-based summary statistics of the WFPT fit with fixed starting point for NCMDT and CMDT and WFPT with fitted starting point for CMDT. * indicates a significance level of <0.05. We used t-tests for all comparisons. Abbreviations: WFPT: Wiener first passage time; NCMDT: Non-cued motion discrimination task; CMDT: Cued motion discrimination task; b: boundary; v: drift; $T_{er}$: non-decision time; $z_0$: starting point; AG: Autism spectrum disorder group; CG: control group.
